# Maternal biomarkers for early prediction of the neural tube defects pregnancies

**DOI:** 10.1101/2020.07.01.20143974

**Authors:** Upendra Yadav, Pradeep Kumar, Vandana Rai

## Abstract

Neural tube defects (NTD) are the most common congenital birth defects. The reason for the NTD cause is still not completely known, but it is believed that some genetic and environmental factors might play a role in its etiology. Among the genetic factors the polymorphism in the folate gene pathway is crucial. Numerous studies have suggested the possible role of maternal higher plasma concentration of homocysteine and low concentration of folate and cobalamin in the development of NTD but some negative studies are also published. So, in this study, authors tried to find out the exact relation between NTD and maternal biomarkers like folate, cobalamin and homocysteine by conducting a meta-analysis. Different electronic databases were searched for the eligible studies. Standardized mean difference (SMD) with 95% confidence interval (CI) was used to determine association between maternal markers as risk for NTD pregnancy. The p value <0.05 was considered statistically significant in all tests. All the statistical analyses were done in the Open Meta-Analyst program. The homocysteine is significantly associated with the increased risk of NTD (SMD= 0.57; 95% CI: 0.35-0.80, p= <0.001; I^2^= 93.01%), s-folate showed protective role in NTD (SMD= −0.48; 95% CI: −0.77 to −0.19, p= 0.001; I^2^= 95.73%), similarly cobalamin is also having protective role (SMD= −0.28; 95% CI: −0.43 to −0.13, p= <0.001; I^2^= 80.40%). In conclusion this study suggest that different maternal biomarkers may be used for the early prediction of the NTDs.

## INTRODUCTION

Neural tube defects (NTD) are very common congenital birth defects [1]. NTD is the general term for a number of malformations but the most common of them are anencephaly, encephalocele and spina bifida. The prevalence of NTD is 1 in 33 infants globally [2]. A recent meta-analysis suggests that the prevalence of NTD in India is 4.5 per 1000 births [3]. NTDs are caused by the failure of closure of neural tube either partially or completely. The reason for the same is still not clear but it is believed that some genetic and environmental factors might play a role in the etiology of NTD [4]. Among the genetic factors the most important is the polymorphism in the methylenetetrahydrofolate reductase (*MTHFR*) gene. This gene has a polymorphism at 677^th^ position which makes this enzyme thermolabile [5]. A recent meta-analysis suggests that polymorphism in this gene increases the chance of the NTD affected pregnancies [6]. The MTHFR enzyme regulates the level of homocysteine. Experimental studies have already suggested that the low level of homocysteine is responsible for the improper closure of the neural tube in mouse model [7]. Several published articles also reported that higher maternal homocysteine concentration is associated with the increased risk of NTD affected pregnancies [8-10]. Higher plasma homocysteine concentration is also reported as to be associated with different diseases like-Down syndrome [11-13], cleft lip and palate [14-16], cardiovascular disease [17-20], diabetes [21], and cancer [22-24] etc.

Although various previous studies have suggested that the elevated level of homocysteine [8-10] or lower level of the folic acid [8, 25, 26] and cobalamin [8, 10, 27] are the risk factors for the etiology of the NTD but the result are conflicting with some negative results. So here in this paper we try to find out the exact relation of these maternal biomarkers with the etiology of the NTDs by conducting a meta-analysis.

## MATERIALS AND METHODS

### Literature search

Different databases (PubMed, ScienceDirect, and SpringerLink) were searched for the eligible studies. The keywords used were “neural tube defects”, or “NTD” in association with “homocysteine”, “folic acid”, or “cobalamin”.

### Inclusion and exclusion criteria

A study included in this meta-analysis only if it was-(i) a case-control study; (ii) reported the level of homocysteine, folic acid, and cobalamin in NTD mothers and control mothers; (iii) either provided mean ± standard deviation (SD) or sufficient data to calculate mean and SD. Similarly, the studies excluded if they were-(i) reviews, meta-analysis, animal model studies, letter to editor, case reports; (ii) not in English.

### Data extraction

From all the eligible articles, following information were extracted-first authors family name; ethnicity; country of study; journals name with year of publication, mean and SD of homocysteine, folic acid and cobalamin. In some publications the authors provided median and range so we calculated the mean and SD as per the method of Hozo et al. [28]. All the information was retrieved by two authors independently (UY and PK) and if any discrepancy found it was sorted out by consultation with the corresponding author.

### Statistical analysis

Standardized mean difference (SMD) with 95% confidence interval (CI) was calculated to determine association between risk for NTD pregnancy and maternal markers i.e. homocysteine, folic acid and cobalamin. The p value <0.05 were considered statistically significant in all tests. The between study heterogeneity was calculated by Cochran’s Q test and quantified by I^2^ tests [29] were applied. If heterogeneity is present (I^2^>50%) random-effects model was applied [30] otherwise fixed-effects model was applied [31]. Publication bias was determined by visualization of the symmetry of the funnel plot. Publication bias was evaluated by the Egger’s linear regression method [32]. All the statistical analyses were done by Open Meta-Analyst program [33]. All p-values were two-tailed with a significance level at 0.05.

## RESULTS

### Characteristics of selected studies and meta-analysis

#### (i) For homocysteine

Five hundred and ninety-six studies were retrieved by electronic database search. Out of which 33 studies were assessed the level of homocysteine in NTD mothers [8-10, 26, 27, 34-61]. Arbour et al. [40] reported two different populations (Cree and non-Cree) we treated them as separate studies so finally for homocysteine we have 34 studies (2,109 cases and 3,514 controls).

Meta-analysis revealed an SMD of 0.57 (95% CI: 0.35-0.80, p= <0.001; I^2^= 93.01%) indicating that elevated level of homocysteine is associated with NTD. High heterogeneity was found so random effect model was applied. Strong correlation was observed in the Asian population (SMD= 1.11, 95% CI: 0.56-1.66, p= <0.001; I^2^= 97.05%). The Caucasian population has low effect of homocysteine (SMD= 0.41, 95% CI: 0.21-0.62, p= <0.001; I^2^= 80.98%). Low effect of homocysteine is also found in the African population (SMD= 0.19, 95% CI: −0.40-0.78, p= 0.52; I^2^= 68.57%) (Table 1; Figure 2).

**Table 1.** Summary estimates for the effect size of different biomarkers in overall and various subgroups, the significance level (p value) of heterogeneity test (Q test), and the I^2^ metric.

| Gene | | Fixed effect<br>SMD (95% CI), p | Random effect<br>SMD (95% CI), p | Heterogeneity<br>p-value (Q<br>test) | $I^2$<br>(%) |
| --- | --- | --- | --- | --- | --- |
| Homocysteine<br>(34 studies) | Overall | 0.34 (0.29-0.40), <0.001 | 0.57 (0.35-0.80), <0.001 | <0.001 | 93.01 |
|  | Asian | 0.40 (0.32-0.49), <0.001 | 1.11 (0.56-1.66), <0.001 | <0.001 | 97.05 |
|  | Caucasian | 0.33 (0.24-0.41), <0.001 | 0.41 (0.21-0.62), <0.001 | <0.001 | 80.98 |
|  | African | 0.11 (-0.19-0.42), 0.47 | 0.19 (-0.40-0.78), 0.52 | 0.07 | 68.57 |
|  | Other | 0.15 (-0.09-0.40), 0.21 | 0.15 (-0.09-0.40), 0.21 | 0.72 | 0 |
| S-folate<br>(36 studies) | Overall | -0.47 (-0.53 to -0.41), <0.001 | -0.48 (-0.77 to -0.19), 0.001 | <0.001 | 95.73 |
|  | Asian | -1.04 (-1.14 to -0.93), <0.001 | -1.37 (-2.14 to -0.61), <0.001 | <0.001 | 97.85 |
|  | Caucasian | -0.23 (-0.31 to -0.15), <0.001 | -0.17 (-0.35 to 0.004), 0.05 | <0.001 | 78.89 |
|  | African | 0.02 (-0.28 to 0.33), 0.87 | -0.03 (-0.56 to 0.49), 0.90 | 0.11 | 60.89 |
|  | Other | 0.12 (-0.08 to 0.34), 0.24 | 0.22 (-0.47 to 0.92), 0.53 | < 0.001 | 89.15 |
| S-cobalamin<br>(29 studies) | Overall | -0.14 (-0.20 to -0.08), <0.001 | -0.28 (-0.43 to -0.13), <0.001 | <0.001 | 80.40 |
|  | Asian | 0.20 (0.08 to 0.31), <0.001 | -0.18 (-0.67 to 0.31), 0.46 | <0.001 | 90.12 |
|  | Caucasian | -0.28 (-0.36 to -0.20), <0.001 | -0.30 (-0.42 to -0.17), <0.001 | 0.004 | 52.95 |
|  | African | -0.35 (-0.66 to -0.04), 0.02 | -0.40 (-0.86 to 0.06), 0.09 | 0.15 | 49.85 |
|  | Other | -0.22 (-0.46 to 0.01), 0.06 | -0.32 (-0.89 to 0.25), 0.27 | 0.02 | 79.48 |

**Figure 1:**
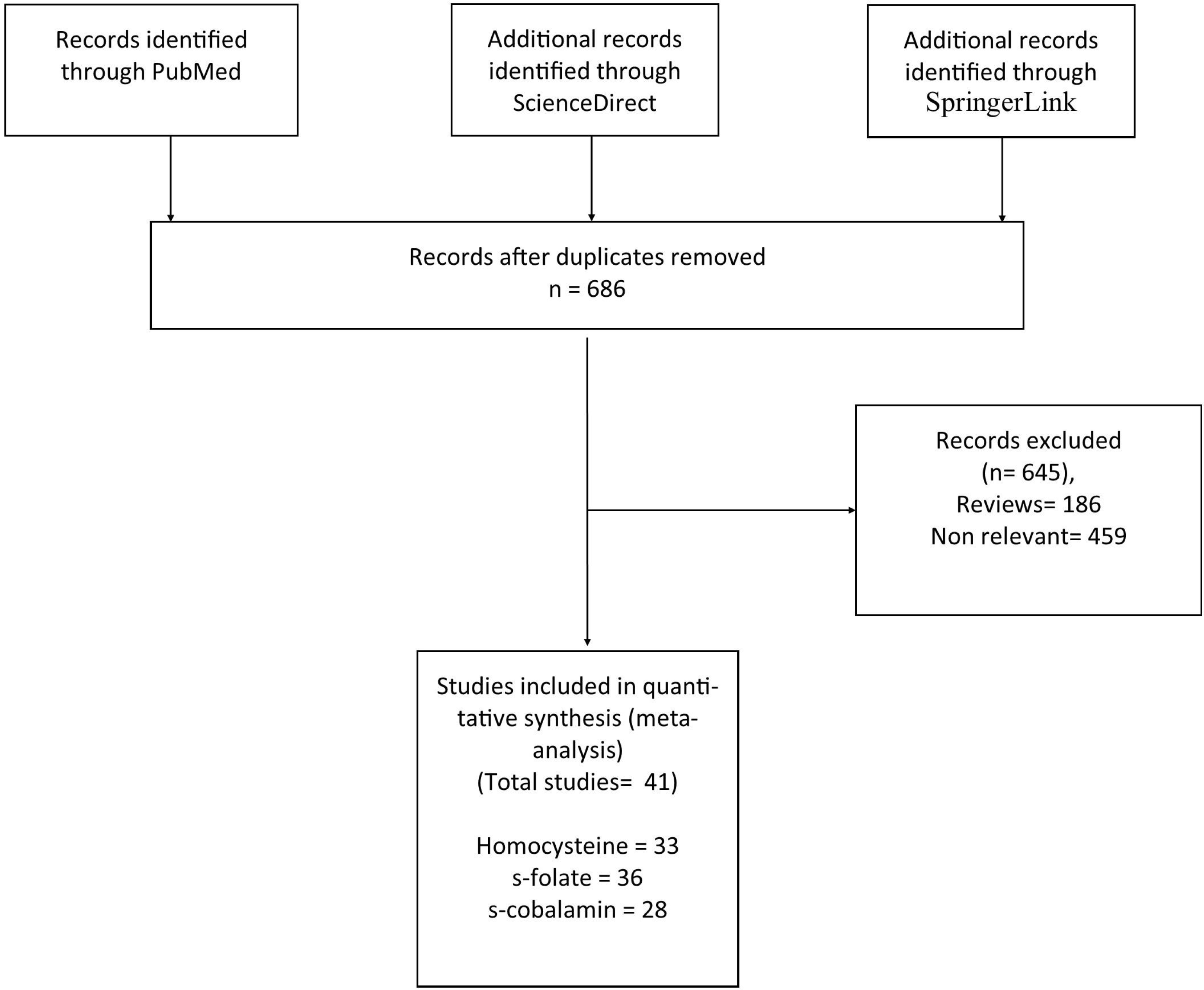
Flow diagram of study search and selection process.

**Figure 2:**
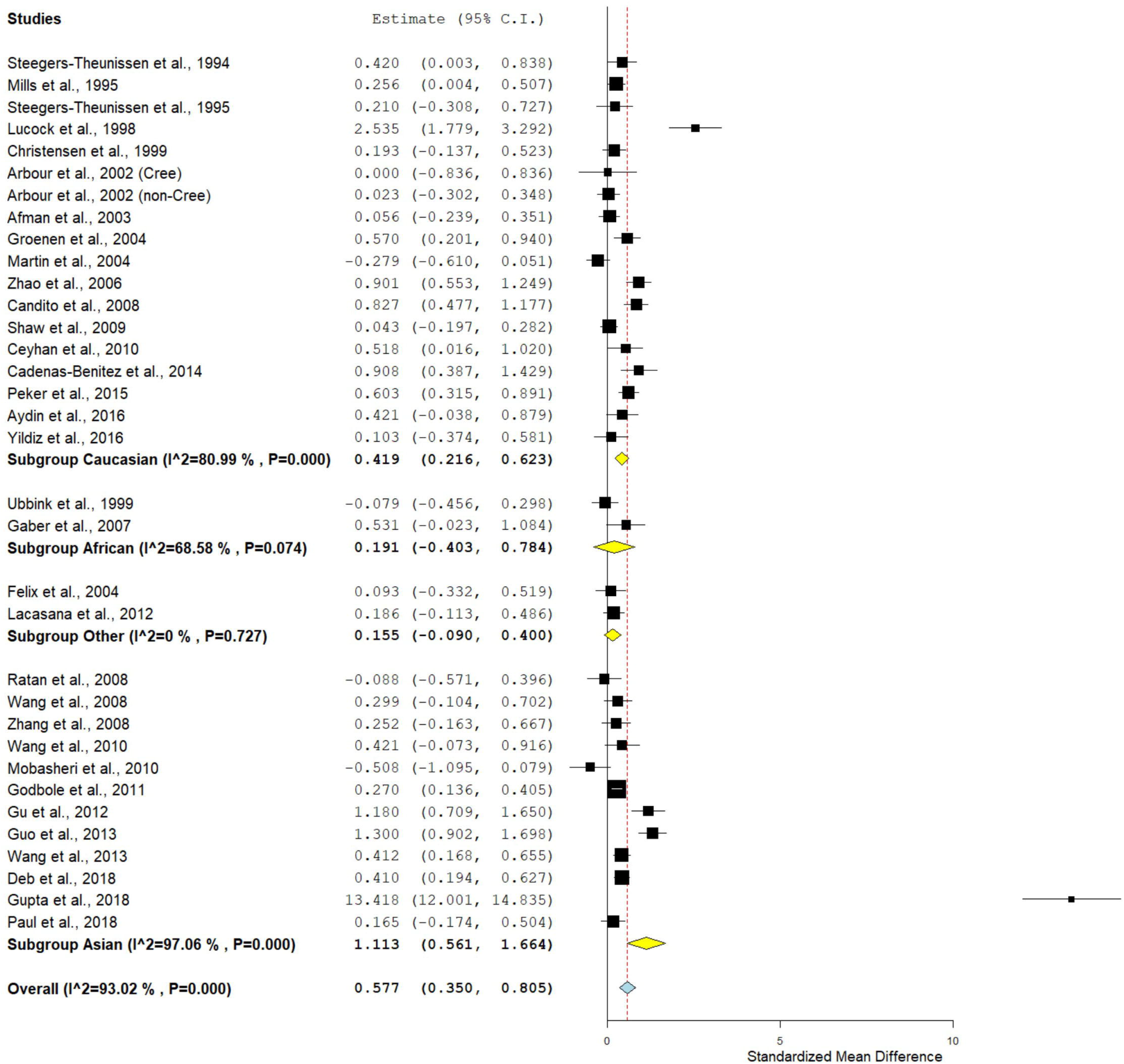
Random effect forest plot of standardized mean differences of homocysteine. Results of individual and mean estimates, and 95% CI of each study were shown. Horizontal lines represented 95% CI, and dotted vertical lines represent the value of the mean.

#### (ii) For s-folate

Three hundred and ninety-three studies were retrieved by our search criteria. Out of which 36 studies were assessed the level of s-folate in NTD mothers [8-10, 25-27, 34-39, 41-43, 45-55, 58, 59, 61-68]. In the selected 36 studies the number of cases and controls were 2,131 and 3,983 respectively.

Meta-analysis revealed an overall SMD of −0.48 (95% CI: −0.77 to −0.19, p= 0.001; I^2^= 95.73%) indicate that elevation in s-folate level in controls play a protective role in the etiology of NTD. High heterogeneity was found so random effect model was applied. The level of s-folate is higher in the Asian population (SMD= −1.37, 95% CI: −2.14 to −0.61, p= <0.001; I^2^= 97.85%) in comparison to the Caucasian population (SMD= −0.17, 95% CI: −0.35 to 0.004, p= 0.05; I^2^= 78.89%). Low effect of s-folate is also found in the African population (SMD= −0.03, 95% CI: −0.56 to 0.49, p= 0.90; I^2^= 60.89%) (Table 1; Figure 3).

**Figure 3:**
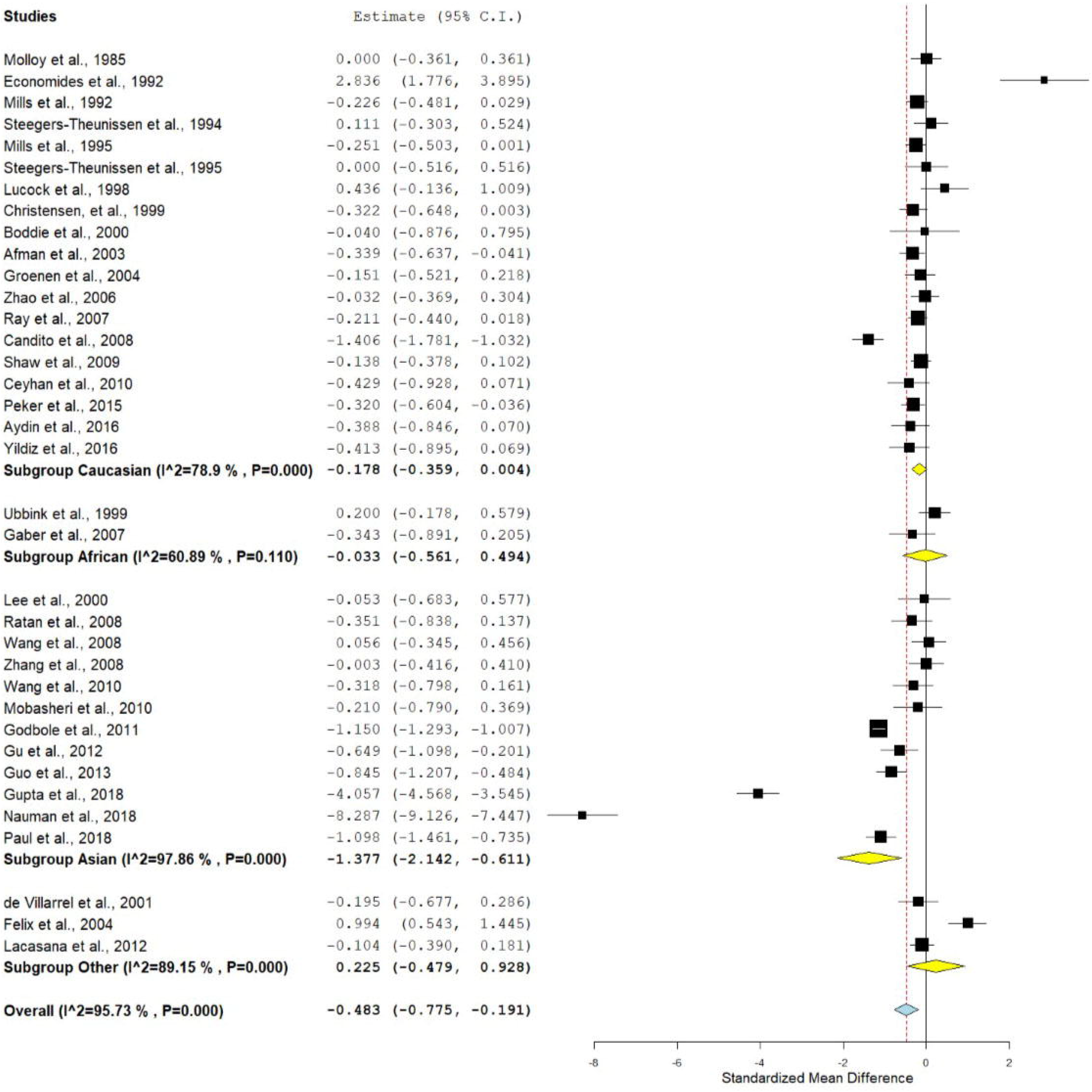
Random effect forest plot of s-RBC folate.

#### (iii) For cobalamin

Three hundred and twenty-six studies were retrieved by our search criteria. Out of which 28 studies were assessed the level of cobalamin in NTD mothers [8, 9, 26, 27, 34-43, 45-47, 49-54, 58, 59, 62-64]. Arbour et al. [40] reported two different populations (Cree and non-Cree) we treated them as separate studies so finally for cobalamin we have 29 studies (1,640 cases and 3,163 controls).

Meta-analysis revealed an overall SMD of −0.28 (95% CI: −0.43 to −0.13, p= <0.001; I^2^= 80.40%) indicate that elevation in cobalamin level in controls shows the decline in the NTD. Higher heterogeneity was found so random effect model was adopted. The level of cobalamin is low in the Asian population (SMD= −0.18, 95% CI: −0.67 to 0.31, p= 0.46; I^2^= 90.12%) in comparison to the Caucasian population (SMD= −0.30, 95% CI: −0.42 to −0.17, p= <0.001; I^2^= 52.95%). Low effect of cobalamin is also found in the African population (SMD= −0.40, 95% CI: −0.86 to 0.06, p= 0.09; I^2^= 49.85%) (Table 1; Figure 4).

**Figure 4:**
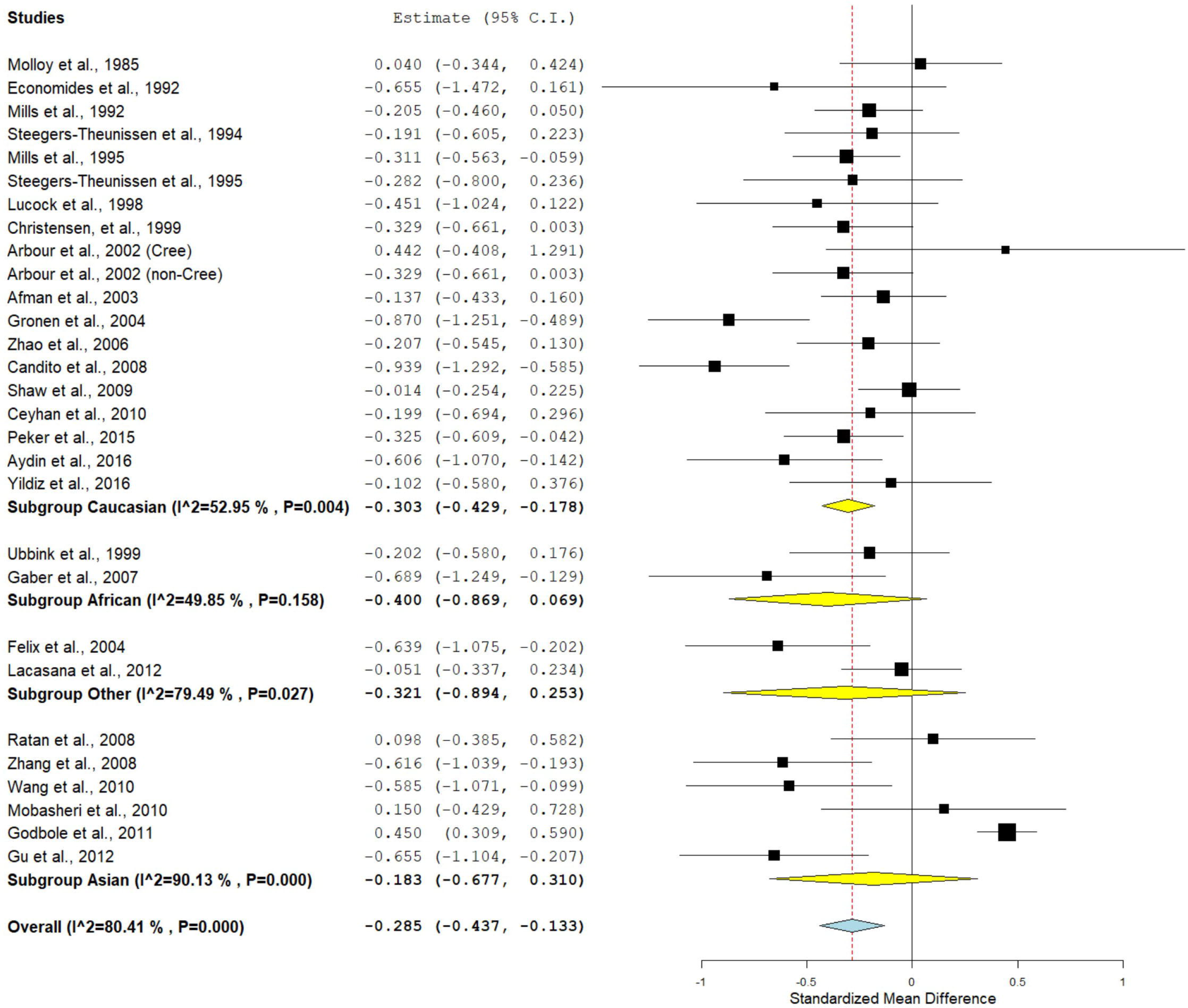
Random effect forest plot of cobalamin.

### Publication bias

The funnel plots were symmetrical in all contrast models (Figure 5), and the P values of Egger’s test were more than 0.05, which provided statistical evidence for the absence of publication bias.

**Figure 5:**
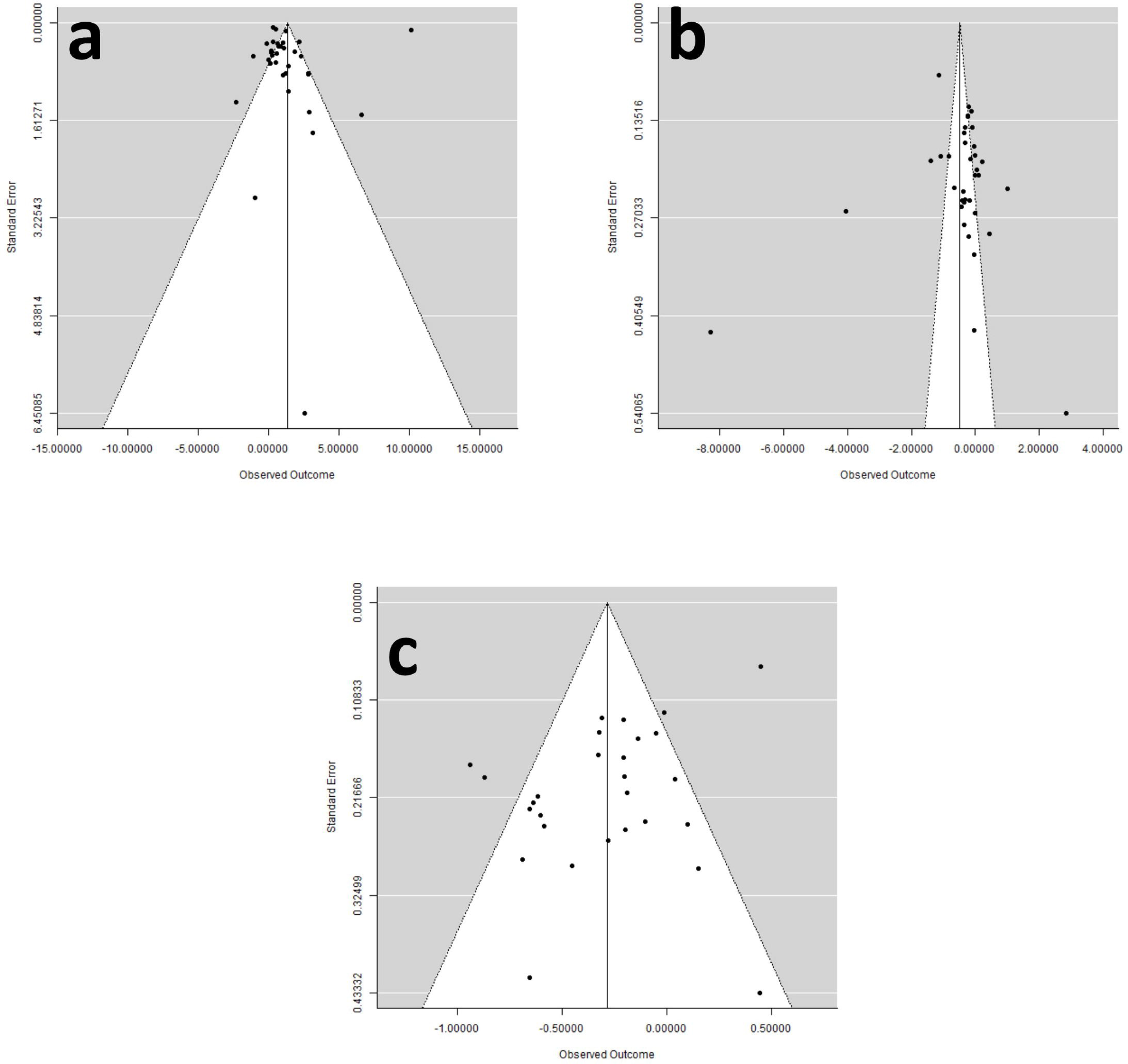
Funnel plots (standard error by mean difference)-a) homocysteine; b) s-RBC folate; c) cobalamin.

## DISCUSSION

This meta-analysis was conducted to check the association between maternal homocysteine, folate and cobalamin levels as risk factors for the etiology of NTDs. We found that the homocysteine is a risk factor for the NTD affected pregnancies. While the folic acid and cobalamin level plays a protective role against the NTD affected pregnancies. It was long known before that the periconceptional folic acid supplementation prevent the NTD [35, 69]. The mechanism by which the NTDs are prevented by dietary folate supplementation is still not fully elucidated. Folic acid is essential for different cellular processes like-cell division, replication and DNA methylation etc. [70-73]. Proper DNA methylation is necessary for the maintenance of chromosome structure and gene expression and both are crucial for the normal development of the fetus [74]. Low plasma folate and /or MTHFR C677T polymorphism increases homocysteine in expecting mothers, which consequently affects the DNA methylation pattern and DNA synthesis of the developing fetus. Improper methylation interferes with the genes regulating neural tube closure [7]. Further higher concentration of homocysteine is toxic and generates free radicals by auto-oxidation and free radicals are toxic for the fetus [75].

Meta-analysis is an effective tool for combining studies having lower effect size. During past decade a number of meta-analyses were published examining the effect of different gene polymorphisms on various disease/disorder susceptibility like-*MTHFR* frequency [76], *MTRR* frequency [77], NTD [6], Down syndrome [78-79], schizophrenia [80-81], epilepsy [82], esophageal cancer [83], breast cancer [84], digestive tract cancer [85], and prostate cancer [86], etc.

During our literature search we found two meta-analyses evaluating the role of homocysteine and NTDs [87, 88]. Tang et al. [87] found that the homocysteine has an association with the NTD with ratio of means (RoM) of 1.16 (95%CI: 1.09-1.23, p= 1.8×10^−6^). They also reported that the NTD-affected mothers have lower levels of folate, RBC folate and vitamin B12. The other study was published in the year 2017 by Yang et al. [88]. The authors of this study only included homocysteine level in their meta-analysis and they found that a significantly higher mean log plasma tHcy level was found in the mothers of NTD affected offspring (log WMD: 0.06; 95%CI: 0.02–0.09, p= 0.001). The present meta-analysis included the larger number of studies with large sample size and results also supports the finding of the previous two studies.

The main strength of our meta-analysis is (i) we included largest sample sizes and also largest number of studies, (ii) we included three parameters *viz*. homocysteine, s-folate, and cobalamin, (iii) we found no publication bias in our study. Here we also want to acknowledge some of the limitations of our meta-analysis such as-(i) use of unadjusted weighted mean difference, (ii) not considered the environmental effect on the metabolites, and (iii) high heterogeneity between studies.

## CONCLUSION

In conclusion, the meat-analysis reveals the role of different biomarkers for the early prediction of the neural tube defects. Also, this study strengths the hypothesis that the elevation in the level of homocysteine or depletion in the level of folate or cobalamin during the pregnancy will increases the chances of an NTD affected offspring.

## Data Availability

All the data are provided in the manuscript.

## ACKNOWLEDGMENTS

Upendra Yadav is highly grateful to VBS Purvanchal University, Jaunpur for providing financial assistance to him in the form of PDF.

## ETHICAL APPROVAL

The article does not contain any studies with human or animal subjects performed by any of the authors.

## REFERENCES

1. Verma IC. High frequency of neural tube defects in North India. Lancet. 1978;1:879–80.

2. WHO Fact sheet No. 370. Congenital Anomalies. October 2012.

3. Allagh KP, Shamanna BR, Murthy GVS, Ness AR, Doyle P, Neogi SB, et al. Birth Prevalence of Neural Tube Defects and Orofacial Clefts in India: A Systematic Review and Meta-Analysis. PLoS ONE. 2015;10(3):e0118961.

4. Wilde JJ, Petersen JR, Niswander L. Genetic, epigenetic, and environmental contributions to neural tube closure. Annu Rev Genet. 2014;48:583–611.

5. Frosst P, Blom HJ, Milos R, Goyette P, Sheppard CA, Mathews RG, et al. A candidate genetic risk factor for vascular disease: a common mutation in methelenetetrahydrofolate reductase. (Letter). Nat Genet. 1995;10:111–3.

6. Yadav U, Kumar P, Yadav SK, Mishra OP, Rai V. Polymorphisms in folate metabolism genes as maternal risk factor for neural tube defects: an updated meta-analysis. Metab Brain Dis. 2015;30:7–24.

7. Blom HJ, Shaw GM, den Heijer M, Finnell RH. Neural tube defects and folate: case far from closed. Nat Rev Neurosci. 2006;7(9):724–31.

8. Candito M, Rivet R, Herbeth B, Boisson C, Rudigoz RC, Luton D, et al. Nutritional and genetic determinants of vitamin B and homocysteine metabolisms in neural tube defects: a multicenter case-control study. Am J Med Genet A. 2008;146A(9):1128–33.

9. Ceyhan ST, Beyan C, Atay V, Yaman H, Alanbay I, Kaptan K, et al. Serum vitamin B12 and homocysteine levels in pregnant women with neural tube defect. Gynecol Endocrinol. 2010;26(8):578–81.

10. Gupta R, Kumari P, Pandey S, Joshi D, Sharma SP, Rai SK, et al. Homocysteine and vitamin B12: Other causes of neural tube defects in Eastern Uttar Pradesh and Western Bihar population. Neurol India. 2018;66(4):1016–1019.

11. James SJ, Pogribna M, Pogribny IP, Melnyk S, Hine RJ, Gibson JB, et al. Abnormal folate metabolism and mutation in the methylenetetrahydrofolate reductase gene may be maternal risk factors for Down syndrome. Am J Clin Nutr. 1999;70(4):495–501.

12. da Silva LR, Vergani N, GaldieriLde C, Ribeiro Porto MP, Longhitano SB, Brunoni D, et al. Relationship between polymorphisms in genes involved in homocysteine metabolism and maternal risk for Down syndrome in Brazil. Am J Med Genet A. 2005;135(3):263–7.

13. Sukla KK, Jaiswal SK, Rai AK, Mishra OP, Gupta V, Kumar A, et al. Role of folate-homocysteine pathway gene polymorphisms and nutritional cofactors in Down syndrome: A triad study. Hum Reprod. 2015;30(8):1982–93.

14. Wong WY, Eskes TK, Kuijpers-Jagtman AM, Spauwen PH, Steegers EA, Thomas CM, et al. Nonsyndromicorofacial clefts: association with maternal hyperhomocysteinemia. Teratology. 1999;60(5):253–7.

15. Blanton SH, Henry RR, Yuan Q, Mulliken JB, Stal S, Finnell RH, et al. Folate pathway and nonsyndromic cleft lip and palate. Birth Defects Res A Clin Mol Teratol. 2011;91(1):50–60.

16. Abdulla R, Tellis RC, Athikari R, Kudkuli J. Evaluation of homocysteine levels in individuals having nonsyndromic cleft lip with or without palate. J Oral Maxillofac Pathol. 2016;20(3):390–394.

17. Wilcken DE, Wilcken B. The pathogenesis of coronary artery disease. A possible role for methionine metabolism. J Clin Invest. 1976;57:1079–82.

18. Boushey CJ, Beresford SAA, Omenn GS, Motulsky AG. A quantitative assessment of plasma homocysteine as a risk factor for vascular disease. JAMA 1995;274:1049–57.

19. Danesh J, Lewington S. Plasma homocysteine and coronary heart disease. Systematic review of published epidemiological studies. J Cardiovasc Risk 1998;5:229–92.

20. Zhang SY, Xuan C, Zhang XC, Zhu J, Yue K, Zhao P, et al. Association Between MTHFR Gene Common Variants, Serum Homocysteine, and Risk of Early-Onset Coronary Artery Disease: A Case-Control Study. Biochem Genet. 2019. doi: 10.1007/s10528-019-09937-x.

21. Huang T, Ren J, Huang J, Li D. Association of homocysteine with type 2 diabetes: a meta-analysis implementing Mendelian randomization approach. BMC Genomics. 2013;14:867.

22. Wu LL, Wu JT. Hyperhomocysteinemia is a risk factor for cancer and a new potential tumor marker. Clin Chim Acta. 2002;322(1-2):21–8.

23. Xu J, Zhao X, Sun S, Ni P, Li C, Ren A, et al. Homocysteine and Digestive Tract Cancer Risk: A Dose-Response Meta-Analysis. J Oncol. 2018;2018:3720684.

24. Hasan T, Arora R, Bansal AK, Bhattacharya R, Sharma GS, Singh LR. Disturbed homocysteine metabolism is associated with cancer. Exp Mol Med. 2019;51(2):21.

25. Martínez de Villarreal LE, Delgado-Enciso I, Valdéz-Leal R, Ortíz-López R, Rojas-Martínez A, Limón-Benavides C, et al. Folate levels and N(5),N(10)-methylenetetrahydrofolate reductase genotype (MTHFR) in mothers of offspring with neural tube defects: a case-control study. Arch Med Res. 2001;32(4):277–82.

26. Aydin H, Arisoy R, Karaman A, Erdogdu E, Çetinkaya A B Geçkinli B, et al. Evaluation of maternal serum folate, vitamin B12, and homocysteine levels and factor V Leiden, factor II g.20210G>A, and MTHFR variations in prenatally diagnosed neural tube defects. Turk J Med Sci. 2016;46(2):489–94.

27. Godbole K, Gayathri P, Ghule S, Sasirekha BV, Kanitkar-Damle A, Memane N, et al. Maternal one-carbon metabolism, MTHFR and TCN2 genotypes and neural tube defects in India. Birth Defects Res A Clin Mol Teratol. 2011;91(9):848–56.

28. Hozo SP, Djulbegovic B, Hozo I. Estimating the mean and variance from the median, range, and the size of a sample. BMC Med Res Methodol. 2005;5:13.

29. Higgins JP, Thompson SG, Deeks JJ, Altman DG. Measuring inconsistency in meta-analyses. BMJ. 2003;327(7414):557–60.

30. DerSimonian R, Laird N. Meta-analysis in clinical trials. Control Clin Trials. 1986;7:177–88.

31. Mantel N, Haenszel W. Statistical aspects of the analysis of data from retrospective studies of disease. J Natl Cancer Inst. 1959;22(4):719–48.

32. Egger M, Davey Smith G, Schneider M, Minder C. Bias in meta-analysis detected by a simple, graphical test. BMJ 1997;315:629–34.

33. Wallace BC, Dahabreh IJ, Trikalinos TA, Lau J, Trow P, Schmid CH. Closing the gap between methodologists and end-users: R as a computational back-end. J Stat Softw. 2013;49:1–15.

34. Steegers-Theunissen RP, Boers GH, Trijbels FJ, Finkelstein JD, Blom HJ, Thomas CM, et al. Maternal hyperhomocysteinemia: a risk factor for neural-tube defects? Metabolism. 1994;43(12):1475–80.

35. Mills JL, McPartlin JM, Kirke PN, Lee YJ, Conley MR, Weir DG, et al. Homocysteine metabolism in pregnancies complicated by neural-tube defects. Lancet. 1995;345(8943):149–51.

36. Steegers-Theunissen RP, Boers GH, Blom HJ, Nijhuis JG, Thomas CM, Borm GF, et al. Neural tube defects and elevated homocysteine levels in amniotic fluid. Am J Obstet Gynecol. 1995;172(5):1436–41.

37. Lucock MD, Daskalakis I, Lumb CH, Schorah CJ, Levene MI. Impaired regeneration of monoglutamyltetrahydrofolate leads to cellular folate depletion in mothers affected by a spina bifida pregnancy. Mol Genet Metab. 1998;65(1):18–30.

38. Christensen B, Arbour L, Tran P, Leclerc D, Sabbaghian N, Platt R, et al. Genetic polymorphisms in methylenetetrahydrofolate reductase and methionine synthase, folate levels in red blood cells, and risk of neural tube defects. Am J Med Genet. 1999;84(2):151–7.

39. Ubbink JB, Christianson A, Bester MJ, Van Allen MI, Venter PA, Delport R, et al. Folate status, homocysteine metabolism, and methylene tetrahydrofolate reductase genotype in rural South African blacks with a history of pregnancy complicated by neural tube defects. Metabolism. 1999;48(2):269–74.

40. Arbour L, Christensen B, Delormier T, Platt R, Gilfix B, Forbes P, et al. Spina bifida, folate metabolism, and dietary folate intake in a Northern Canadian aboriginal population. Int J Circumpolar Health. 2002;61(4):341–51.

41. Afman LA, Blom HJ, Van der Put NM, Van Straaten HW. Homocysteine interference in neurulation: a chick embryo model. Birth Defects Res A Clin Mol Teratol. 2003;67(6):421–8.

42. Félix TM, Leistner S, Giugliani R. Metabolic effects and the methylenetetrahydrofolate reductase (MTHFR) polymorphism associated with neural tube defects in southern Brazil. Birth Defects Res A Clin Mol Teratol. 2004;70(7):459–63.

43. Groenen PM, van Rooij IA, Peer PG, Gooskens RH, Zielhuis GA, Steegers-Theunissen RP. Marginal maternal vitamin B12 status increases the risk of offspring with spina bifida. Am J Obstet Gynecol. 2004;191(1):11–7.

44. Martín I, Gibert MJ, Pintos C, Noguera A, Besalduch A, Obrador A. Oxidative stress in mothers who have conceived fetus with neural tube defects: the role of aminothiols and selenium. Clin Nutr. 2004;23(4):507–14.

45. Zhao W, Mosley BS, Cleves MA, Melnyk S, James SJ, Hobbs CA. Neural tube defects and maternal biomarkers of folate, homocysteine, and glutathione metabolism. Birth Defects Res A Clin Mol Teratol. 2006;76(4):230–6.

46. Gaber KR, Farag MK, Soliman SE, El-Bassyouni HT, El-Kamah G. Maternal vitamin B12 and the risk of fetal neural tube defects in Egyptian patients. Clin Lab. 2007;53(1-2):69–75.

47. Ratan SK, Rattan KN, Pandey RM, Singhal S, Kharab S, Bala M, et al. Evaluation of the levels of folate, vitamin B12, homocysteine and fluoride in the parents and the affected neonates with neural tube defect and their matched controls. Pediatr Surg Int. 2008;24(7):803–8.

48. Wang Y, Zhang HY, Liang QL, Yang HH, Wang YM, Liu QF, et al. Simultaneous quantification of 11 pivotal metabolites in neural tube defects by HPLC-electrospray tandem mass spectrometry. J Chromatogr B Analyt Technol Biomed Life Sci. 2008;863(1):94–100.

49. Zhang HY, Luo GA, Liang QL, Wang Y, Yang HH, Wang YM, et al. Neural tube defects and disturbed maternal folate- and homocysteine-mediated one-carbon metabolism. Exp Neurol. 2008;212(2):515–21.

50. Shaw GM, Finnell RH, Blom HJ, Carmichael SL, Vollset SE, Yang W, et al. Choline and risk of neural tube defects in a folate-fortified population. Epidemiology. 2009;20(5):714–9.

51. Wang L, Wang F, Guan J, Le J, Wu L, Zou J, et al. Relation between hypomethylation of long interspersed nucleotide elements and risk of neural tube defects. Am J Clin Nutr. 2010;91(5):1359–67.

52. Mobasheri E, Keshtkar A, Golalipour MJ. Maternal folate and vitamin b(12) status and neural tube defects in northern Iran: a case control study. Iran J Pediatr. 2010;20(2):167–73.

53. Gu Q, Li Y, Cui ZL, Luo XP. Homocysteine, folate, vitamin B12 and B6 in mothers of children with neural tube defects in Xinjiang, China. Acta Paediatr. 2012;101(11):e486–90.

54. Lacasaña M, Blanco-Muñoz J, Borja-Aburto VH, Aguilar-Garduño C, Rodríguez-Barranco M, Sierra-Ramirez JA, et al. Effect on risk of anencephaly of gene-nutrient interactions between methylenetetrahydrofolate reductase C677T polymorphism and maternal folate, vitamin B12 and homocysteine profile. Public Health Nutr. 2012;15(8):1419–28.

55. Guo J, Xie H, Wang J, Zhao H, Wang F, Liu C, et al. The maternal folate hydrolase gene polymorphism is associated with neural tube defects in a high-risk Chinese population. Genes Nutr. 2013;8(2):191–7.

56. Wang F, Wang J, Guo J, Chen X, Guan Z, Zhao H, et al. PCMT1 gene polymorphisms, maternal folate metabolism, and neural tube defects: a case-control study in a population with relatively low folate intake. Genes Nutr. 2013;8(6):581–7.

57. Cadenas-Benitez NM, Yanes-Sosa F, Gonzalez-Meneses A, Cerrillos L, Acosta D, Praena-Fernandez JM, et al. Association of neural tube defects in children of mothers with MTHFR 677TT genotype and abnormal carbohydrate metabolism risk: a case-control study. Genet Mol Res. 2014;13(1):2200–7.

58. Peker E, Demir N, Tuncer O, Üstyol L, Balahoroğlu R, Kaba S, et al. The levels of vitamin B12, folate and homocysteine in mothers and their babies with neural tube defects. J Matern Fetal Neonatal Med. 2016;29(18):2944–8.

59. Yildiz SH, OzdemirErdogan M, Solak M, Eser O, ArıkanTerzi ES, Eser B, et al. Lack of association between the methylenetetrahydropholatereductase gene A1298C polymorphism and neural tube defects in a Turkish study group. Genet Mol Res. 2016;15(2):gmr.15028051.

60. Deb R, Arora J, Samtani R, Garg G, Saksena D, Sharma N, et al. Folic acid, dietary habits, and homocysteine levels in relation to neural tube defects: A case-control study in North India. Birth Defects Res. 2018;110(14):1148–1152.

61. Paul S, Sadhukhan S, Munian D, Bankura B, Das M. Association of FOLH1, DHFR, and MTHFR gene polymorphisms with susceptibility of Neural Tube Defects: A case control study from Eastern India. Birth Defects Res. 2018;110(14):1129–1138.

62. Molloy AM, Kirke P, Hillary I, Weir DG, Scott JM. Maternal serum folate and vitamin B12 concentrations in pregnancies associated with neural tube defects. Arch Dis Child. 1985;60(7):660–5.

63. Economides DL, Ferguson J, Mackenzie IZ, Darley J, Ware II, Holmes-Siedle M. Folate and vitamin B12 concentrations in maternal and fetal blood, and amniotic fluid in second trimester pregnancies complicated by neural tube defects. Br J Obstet Gynaecol. 1992;99(1):23–5.

64. Mills JL, Tuomilehto J, Yu KF, Colman N, Blaner WS, Koskela P, et al. Maternal vitamin levels during pregnancies producing infants with neural tube defects. J Pediatr. 1992;120(6):863–71.

65. Boddie AM, Dedlow ER, Nackashi JA, Opalko FJ, Kauwell GP, Gregory JF 3rd, et al. Folate absorption in women with a history of neural tube defect-affected pregnancy. Am J Clin Nutr. 2000;72(1):154–8.

66. Lee BH, Cheong HI, Shin YS, Cho BK, Wang KC. The effect of C677T mutation of methylene tetrahydrofolate reductase gene and plasma folate level on hyperhomocysteinemia in patients with meningomyelocele. Childs Nerv Syst. 2000;16(9):559–63.

67. Ray JG, Wyatt PR, Thompson MD, Vermeulen MJ, Meier C, Wong PY, et al. Vitamin B12 and the risk of neural tube defects in a folic-acid-fortified population. Epidemiology. 2007;18(3):362–6.

68. Nauman N, Jalali S, Shami S, Rafiq S, Große G, Hilger AC, et al. Low maternal folate concentrations and maternal MTHFR C677T polymorphism are associated with an increased risk for neural tube defects in offspring: a case-control study among Pakistani case and control mothers. Asia Pac J ClinNutr. 2018;27(1):253–260.

69. Kirke PN, Molloy AM, Daly LE, Burke H, Weir DG, Scott JM. Maternal plasma folate and vitamin B12 are independent risk factors for neural tube defects. QJ Med. 1993;86:703–8.

70. Blount BC, Mack MM, Wehr CM, MacGregor JT, Hiatt RA, Wang G, et al. Folate deficiency causes uracil misincorporation into human DNA and chromosome breakage: implications for cancer and neuronal damage. Proc Natl Acad Sci USA. 1997;94:3290–5.

71. Morrison K, Papapetrou C, Hol FA, Mariman EC, Lynch SA, Burn J, et al. Susceptibility to spina bifida; an association study of five candidate genes. Ann Hum Genet. 1998;62(Pt5):379–96.

72. James SJ, Pogribny IP, Pogribna M, Miller BJ, Jernigan S, Melnyk S. Mechanisms of DNA damage, DNA hypomethylation, and tumor progression in the folate/methyl-deficient rat model of hepatocarcinogenesis. J Nutr. 2003;133(11Suppl.1):37.

73. Pogribny IP, James SJ, Jernigan S, Pogribna M. Genomic hypomethylation is specific for preneoplastic liver in folate/methyl deficient rats and does not occur in non-target tissues. Mutat Res. 2004;548(1-2):53–9.

74. Razin A, Kantor B. DNA methylation in epigenetic control of gene expression. Prog Mol Subcell Biol. 2005;38:151–67.

75. Tyagi N, Sedoris KC, Steed M, Ovechkin AV, Moshal KS, Tyagi SC. Mechanisms of homocysteine-induced oxidative stress. Am J Physiol Heart Circ Physiol. 2005;289:H2649–56.

76. Yadav U, Kumar P, Gupta S, Rai V. Distribution of MTHFR C677T Gene Polymorphism in Healthy North Indian Population and an Updated Meta-analysis. Indian J Clin Biochem. 2017;32(4):399–410.

77. Yadav U, Kumar P, Rai V. Distribution of Methionine Synthase Reductase (MTRR) Gene A66G Polymorphism in Indian Population. Ind J Clin Biochem (2019). https://doi.org/10.1007/s12291-019-00862-9.

78. Rai V, Yadav U, Kumar P, Yadav SK, Mishra OP. Maternal methylenetetrahydrofolate reductase C677T polymorphism and Down syndrome risk: a meta-analysis from 34 studies. Plos One. 2014;9(9):e108552.

79. Rai V, Yadav U, Kumar P. Null association of maternal MTHFR A1298C polymorphism with Down syndrome pregnancy: An updated meta-analysis. Egypt J Med Hum Genet. 2017;18:9–18.

80. Yadav U, Kumar P, Gupta S, Rai V. Role of MTHFR C677T gene polymorphism in the susceptibility of schizophrenia: an updated meta-analysis. Asian J Psychiatr. 2016;20:41–51.

81. Rai V, Yadav U, Kumar P, Yadav SK, Gupta S. Methylenetetrahydrofolate reductase A1298C genetic variant & risk of schizophrenia: A meta-analysis. Indian J Med Res. 2017;145(4):437–47.

82. Rai V, Kumar P. Methylenetetrahydrofolate reductase C677T polymorphism and susceptibility to epilepsy. Neurol Sci. 2018;39(12):2033–41.

83. Kumar P, Rai V. Methylenetetrahydrofolate reductase C677T polymorphism and risk of esophageal cancer: An updated meta-analysis. Egypt J Med Hum Genet. 2018;19(4): 273–84.

84. Rai V, Yadav U, Kumar P. Impact of Catechol-O-Methyltransferase Val 158Met (rs4680) Polymorphism on Breast Cancer Susceptibility in Asian Population. Asian Pac J Cancer Prev. 2017;18(5):1243–50.

85. Yadav U, Kumar P, Rai V. NQO1 Gene C609T Polymorphism (dbSNP: rs1800566) and Digestive Tract Cancer Risk: A Meta-Analysis. Nutr Cancer. 2018;70(4):557–68.

86. Yadav U, Kumar P, Rai V. Role of MTHFR A1298C gene polymorphism in the etiology of Prostate cancer: A systematic review and updated meta-analysis. Egypt J Med Hum Genet. 2016;17:141–8.

87. Tang KF, Li YL, Wang HY. Quantitative assessment of maternal biomarkers related to one-carbon metabolism and neural tube defects. Sci Rep. 2015;5:8510.

88. Yang M, Li W, Wan Z, Du Y. Elevated homocysteine levels in mothers with neural tube defects: a systematic review and meta-analysis. J Matern Fetal Neonatal Med. 2017;30(17):2051–7.

